# Pre-COVID-19 humoral immunity to common coronaviruses does not confer cross-protection against SARS-CoV-2

**DOI:** 10.1101/2020.08.14.20173393

**Authors:** Makoto Miyara, Delphine Sterlin, François Anna, Stéphane Marot, Alexis Mathian, Mo Atif, Paul Quentric, Audrey Mohr, Laetitia Claër, Christophe Parizot, Karim Dorgham, Hans Yssel, Thibaud Chazal, Jehane Fadlallah, Julien Haroche, Neila Benameur, David Boutolleau, Sonia Burrel, Sasi Mudumba, Rick Hockett, Cary Gunn, Pierre Charneau, Vincent Calvez, Anne-Geneviève Marcellin, Zahir Amoura, Guy Gorochov

**Affiliations:** Sorbonne Université, Inserm, Centre d’Immunologie et des Maladies Infectieuses (CIMI-Paris), Assistance Publique Hôpitaux de Paris (AP-HP), Hôpital Pitié-Salpêtrière, 75013 Paris, France; Unit of Antibodies in Therapy and Pathology, Institut Pasteur, UMR1222 Inserm, 75015 Paris, France; Unité de Virologie Moléculaire et Vaccinologie, Institut Pasteur, 25-28 Rue du Dr Roux, 75015 Paris, France. Decaler vers le haut; Theravectys, France; Sorbonne Université, Inserm, Institut Pierre Louis d’Epidémiologie et de Santé Publique (iPLESP), AP-HP, Hôpital Pitié Salpêtrière, Service de Virologie, Paris, France; Service de Médecine Interne 2, Institut E3M, Assistance Publique Hôpitaux de Paris (AP-HP), Hôpital Pitié-Salpêtrière, 75013 Paris, France; Genalyte Inc., San Diego, CA, United States of America

## Abstract

It is currently unknown whether acquired immunity to common alpha- and beta-coronaviruses provides cross-protection against SARS-CoV-2. In this study, we found that certain patient sera and intravenous immunoglobulins (IVIG) collected prior to the COVID-19 outbreak were cross-reactive to SARS-CoV-2 full-length Spike, S2 domain, and Nucleocapsid. However, their presence did not translate into neutralizing activity against SARS-CoV-2 *in vitro*. Importantly, we detected serum IgG reactivity to common coronaviruses in the early sera of patients with severe COVID-19 before the appearance of anti-SARS-CoV-2 antibodies. Collectively, the results of our study indicate that pre-existing immunity to common coronaviruses does not confer cross-protection against SARS-CoV-2 *in vivo*.

## Main

The ongoing pandemic related to SARS-CoV2 has demonstrated significant heterogeneity across global populations, as well as differing levels of infection severity^1^. In addition to these unexplained variations, children and young adults especially seem to be better protected from the severe form of coronavirus disease 2019 (COVID-19). For this reason, it has been hypothesised that any past infections or immunisations related to the common alpha-coronavirus HCoV-NL-63 and -229-E and beta-coronavirus HCoV-OC-43 and -HK-U1 could lead to cross-protection against SARS-CoV-2^2-5^, although cross-neutralising antibody responses were until now never reported.

In order to test this hypothesis, we first analysed the sera of 76 healthy French donors (48 males; 28 females; median age of 39 years; age range 19-65, **supplementary table 1**) drawn in 2015. Although we did not detect anti-Receptor Binding Domain (RBD) reactivity in these European individuals, we found that six serological samples (7.9%) were reactive to one or several of the others SARS-COV2 antigens (S2 domain, full-length Spike, and/or Nucleocapsid). Moreover, these sera also randomly recognized all common coronaviruses, which indicated that pre-COVID cross-reactivity to SARS-CoV-2 antigens was neither specific to a unique family nor a type of coronavirus **(Figure 1a)**. From a humoral immunity perspective, we also assessed the immunoglobulin G (IgG) reactivity of intravenous immunoglobulins (IVIG) manufactured prior to the COVID-19 outbreak. This was important because therapeutic IVIG consists of IgG isolated from about 10 000 pooled plasma samples of healthy donors. Hence, these products could be a useful indicator of any pre-existing humoral response of the general population^6^. Our investigation into the IgG reactivity of three different batches of IVIG demonstrated high reactivity towards all common coronaviruses, as well as detectable reactivity to the SARS-CoV-2 S2 domain and full-length Spike but not to Nucleocapsid **(Figure 1b)**. Collectively, the results indicate that pre-COVID cross-reactivity to SARS-CoV-2 antigens is not rare in the general population.

Furthermore, to determine whether such cross-reactivity was bidirectional (i.e. originating from SARS-CoV-2 but reacting to common coronaviruses too), we sequentially analysed the sera of eight severe COVID-19 patients (clinical features in **Supplementary Table 2**) for their IgG reactivity to SARS-CoV-2 and common coronaviruses (**Figure 1c**). Indeed, we observed a parallel increase in IgG antibody titers directed towards SARS-CoV-2 antigens and the beta-coronaviruses HCoV-OC-43 and HCoV-HK-U1. However, no increase in responses to alpha-coronavirus was identified. These IgG responses to common beta-coronaviruses were more likely due to cross-reactivity and not because of ongoing infection related to other coronaviruses because the nasopharyngeal RT-PCR of our patients was specifically positive for SARS-CoV-2 and negative for all other common coronaviruses (**Supplementary Table 2**). Moreover, the early sera from all patients but one showed reactivity to beta-coronaviruses HCoV-OC-43 and HCoV-HK-U1, whereas the early sera from two patients also recognized alpha-coronaviruses HCoV-NL-63 and HCoV-229-E. Importantly, these responses were present before the appearance of anti-SARS-CoV-2 antibodies (**Figure 1c**). Collectively, these data indicate that pre-existing immunity to common coronaviruses did not prevent the onset of COVID-19 in these patients.

**Figure 1.**
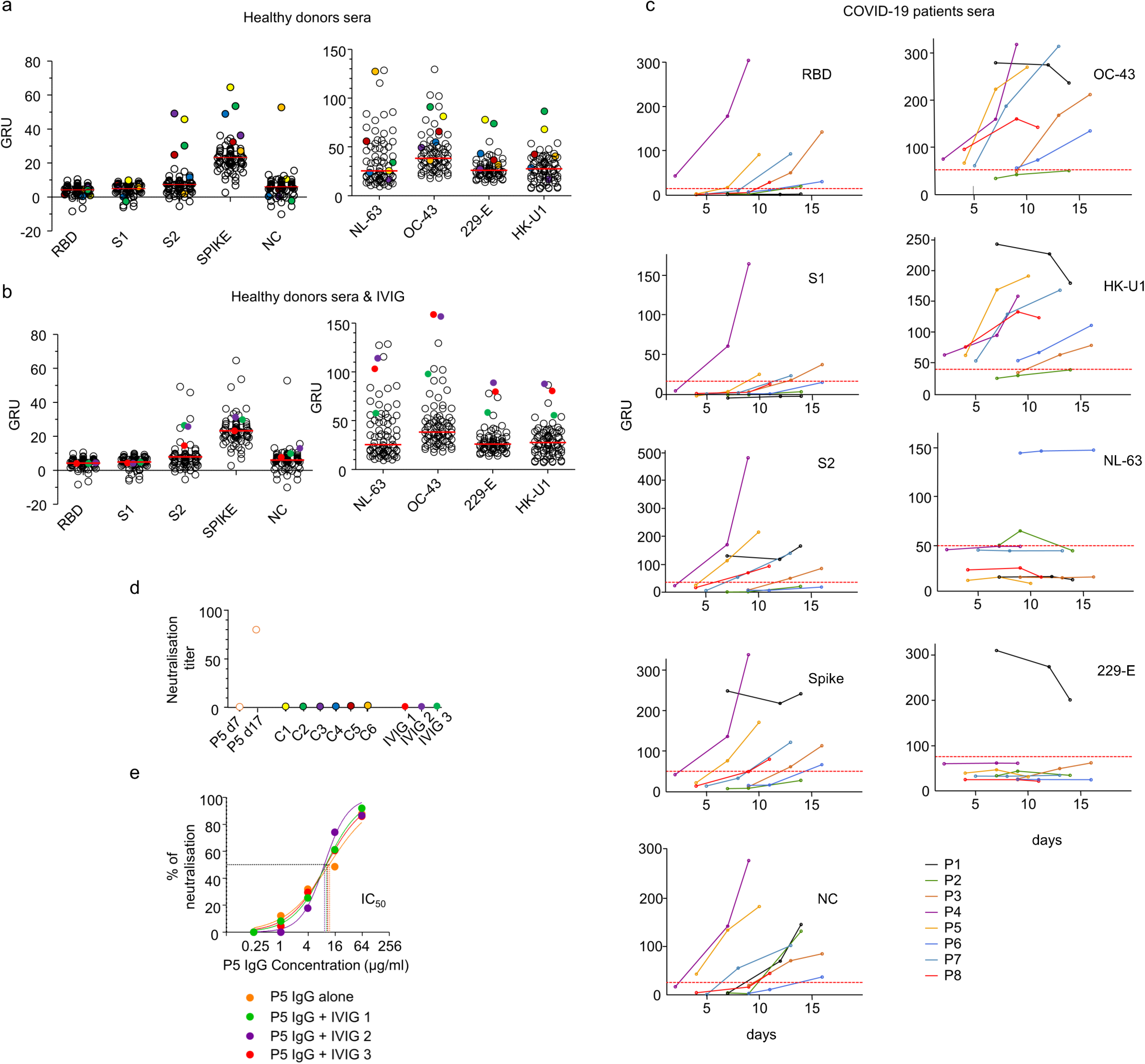
(a) IgG reactivity of 76 sera drawn from healthy donors in 2015 analysed by phototonic ring immunoassay for SARS-CoV-2 antigens: Receptor binding domain (RBD), S1 domain (S1), S2 domain (S2) of Spike protein, Spike and Nucleocapsid (NC), (left) and to common coronaviruses HCoV-OC-43, -229-E and -HK-U1 spike proteins and HCoV-NL-63 Nucleoprotein (right). 6 sera (C1 to C6) positive for either SARS-CoV-2 S2, Spike or NC are show in circled colored dots. Reactivity levels are reported in GRU (Genalyte reactive units). Median reactivities are shown with red horizontal lines. (b) IgG reactivity to SARS-CoV-2 and to common coronaviruses antigens of 3 batches of IVIG produced before the outbreak of COVID-19 and of the 76 sera described in (a). IVIG batches (IVIG 1 to 3) are shown in uncircled colored dots. Antigens are described in (a). Reactivity levels are reported in GRU (Genalyte reactive units). (c) Time course of IgG reactivity to SARS-CoV-2 antigens and to common coronaviruses of the sera of 8 patients (P1 to P8) with confirmed severe COVID-19. Dotted red lines indicate threshold for positivity. (d) Neutralisation capacities of pre-COVID sera cross-reactive to SARS-CoV-2 antigens, of IVIG batches and of the sera of Patient 5 at day 7 (P5 d7) and day 17 (P5 d17) after the onset of the symptoms. Neutralisation antibody titers are expressed as the highest serum dilution which shows 100% inhibition of the cytopathic effect of live SARS-CoV-2 on Vero E6 cells. (e) Neutralisation capacities on pseudotyped vectors of the IgG isolated from the serum of COVID19 Patient 5 drawn 17 days after symptom onset, diluted with standard diluant (orange) or with IVIG batches (green, purple and red) with a final IVIG IgG concentration of 12 mg/mL in the neutralisation assay. Half maximal inhibitory concentrations IC_50_ of P5 serum alone (11.67 μg/mL) or diluted with IVIG1 (9.94 μg/mL), IVIG2 (9.14 μg/mL) and IVIG3 (10.15 μmg/mL) are shown with dotted lines.

In order to experimentally confirm that pre-existing cross-reactivity did not lead to cross-protection against SARS-CoV-2 infection, we assessed the neutralising capacities of the six pre-COVID-19 sera reactive for SARS-CoV-2 antigens and of the three IVIG batches using an *in vitro* whole SARS-CoV-2 neutralization assay. In COVID-19, whilst full neutralisation of SARS-CoV-2 is observed with sera containing anti-RBD antibodies, none is observed using earlier serum devoid of detectable anti-RBD antibodies^7,8^.

This effect is best illustrated in **Figure 1d** with sera of Patient 5 being able to neutralise SARS-CoV-2 when drawn 17 days after symptom onset but not at day 7. In addition, we found that none of the pre-COVID-19 sera, nor IVIG batches were able to neutralise the replication of SARS-CoV-2 *in vitro*. To ensure that no inhibitor, possibly introduced during the IVIG manufacturing process, had interfered with any neutralising IgG present in these preparations, we assessed the neutralising capacities of IgG isolated from Patient 5 diluted with increasing volumes of IVIG. As shown in **Figure 1e**, purified IgG from Patient 5 were able to potently neutralise even when diluted with IVIG. Indeed, the half-maximal inhibitory concentrations IC_50_ of Patient 5 serum alone (11.67 μg/mL) or diluted with IVIG (IVIG1: 9.94 μg/mL, IVIG2: 9.14 μg/mL, IVIG3: 10.15 μg/mL) were similar. Collectively, these results confirm that IVIG products manufactured before the COVID-19 outbreak have no neutralising capabilities and that, on the contrary, the addition of neutralising IgG from COVID-19 patients to IVIG preparations could confer such neutralising capabilities. Since about 10 μg/mL of COVID-19 patient IgG are sufficient to neutralise 50% of the viral cytopathic effect and 64 μg/mL to neutralise all of the viral cytopathic effect in the presence of IVIG (**Figure 1e**), and since IVIG were tested for neutralisation at 12 mg/mL, we can estimate that the presence of only 1 SARS-CoV-2-immunized individual out of 1200 donors (0.08 %) would have been sufficient to confer detectable anti-SARS-CoV-2 activity, while one such immunized individual out of 187.5 donors (0.5%) would have conferred potent neutralising capacities to IVIG.

Overall, our results indicate that cross-reactivity does occur between common coronaviruses and SARS-CoV-2. Cross-protection to SARS-CoV-2 has already been observed with sera isolated from patients previously infected by SARS-CoV^9^. This cross-protection is well explained by the high homology between the SARS-CoV and SARS-CoV-2 RBD. Whilst there are several homologous regions in the S2 domain between SARS-CoV-2 and common alpha- and beta-coronaviruses, that could account for the observed cross-reactivity, there is no homology between SARS-CoV-2 and common alpha- and beta-coronaviruses in the RBD regions^4,10,11^. Indeed, antibodies to the RBDs of common coronaviruses are commonly detected in most adults but do not cross-react with the SARS-CoV-2 RBD^8^. In this report, we confirm that pre-COVID-19 immunity to common coronaviruses does not confer cross-protection against SARS-CoV-2. Variations between common coronaviruses and SARS-CoV-2 RBDs are likely to be the reason why we did not observe cross-protection from common alpha-and beta-coronavirus immunity against SARS-CoV-2 *in vivo* and *in vitro*.

Finally, we also demonstrated that IVIG batches produced prior to the outbreak of COVID-19 do not have neutralising capacities. Importantly, these preparations also did not interfere with the SARS-CoV-2 neutralising capacity of serum IgG. This result suggests that IVIG batches manufactured after the COVID-19 outbreak should not exclude donors that have recovered from COVID-19, provided that they would not present potentially deleterious anti-self reactivities or antibody-dependent SARS-CoV-2 enhancement activity^12^. It is nevertheless important to consider that IVIG infusions were ineffective in non-COVID-19-related SARS^13^. Similarly, no efficacy was yet demonstrated in treating COVID-19 patients using plasma obtained from those that had recovered from the infection^14^. Hence, we consider that IVIG batches manufactured during the current pandemic are unlikely to perform a curative role. It will be nevertheless important to assess “post-COVID” IVIG as a prophylactic, or as a preventive treatment in early COVID-19 infections instead.

## Data Availability

All raw data are available in supplementary data or upon request

## Materials & Methods

### Patient and healthy donor samples and IVIG batches

Collection of blood from anonymous healthy donors was carried out at the French Institute of Blood Donation (EFS, Etablissement français du sang, Paris, France) in 2015 in line with Local and Regional Ethics Committee “CPP - Ile de France-VI” at the Pitié-Salpêtrière Hospital. The patient samples were collected from 8 COVID-19 patients referred to the Department of Internal Medicine 2 at the Pitié-Salpêtrière Hospital, Paris. Demographic and clinical characteristics are detailed in **Supplementary Table 2**. All patients or their relatives gave informed consent. This study was approved by the Local Ethical Committee of Sorbonne Université (n°2020-CER2020-21). For all samples, sera were stored immediately after collection at −80°C. Three batches of IVIG pharmaceutical products (TEGELINE, LFB; CLAIRYG, LFB and PRIVIGEN, CSL BEHRING), manufactured before 2019 in France, were assessed with a 12mg/mL IgG concentration.

### Photonic ring immunoassay

SARS-CoV-2 specific IgG antibodies were measured with The Maverick TM SARS-CoV-2 Multi-Antigen Serology Panel (Genalyte Inc. USA) according to the manufacturer’s instructions. The Maverick TM SARS-CoV-2 Multi-Antigen Serology Panel (Genalyte Inc) is designed to detect antibodies to five SARS-CoV-2 antigens; nucleocapsid, Spike S1 RBD, Spike S1S2, Spike S2, and Spike S1, and common coronavirus HCoV-NL-63 Nucleocapsid, HCoV-OC-43, HCoV-229-E and HCoV-HK-U1 Spike proteins within a multiplex format based on photonic ring resonance technology^15,16^. This system detects and measures changes in resonance when antibodies bind to their respective antigens on the chip. Threshold values for positivity were set by the manufacturer. Raw data are available in **Supplementary Tables 1 and 3**.

### Whole virus neutralisation test (VNT)

Neutralising activity of the different sera was tested with a whole virus replication assay for which we used a SARS-CoV-2 strain isolated from a COVID-19 patient at the Pitié-Salpêtrière Hospital, Paris. This patient was confirmed positive by a SARS-CoV-2 RT-PCR on March 6^th^, 2020, and the virus was isolated by inoculating Vero cells with a sputum sample in our biosafety level-3 (BSL-3) facility. The serum samples were decomplemented by heat inactivation (56°C for 30 min), 2-fold serially diluted (starting at 1:5 to 1:2560), and then pre-incubated on a 96-well plate with 50 μl of diluted virus (2.10^3^ Fifty percent Tissue Culture Infective Dose TCID50/mL) at 37°C for 60 min. Next, 100 μL of Vero E6 cells suspension (3.10^5^ cells/mL) was added to the mixture and incubated at 37°C with 5% CO_2_ until a microscopic examination was performed on day 4 for the cytopathic effect (CPE). Neutralisation antibody titers were expressed as the highest serum dilution that showed 100% inhibition of CPE. A same positive serum was added to each experiment to assess the reproducibility of the VNT.

### Purification of IgG from serum

IgG were isolated from serum samples diluted in 1X-PBS as previously described^17^. Briefly, serum samples were loaded onto Protein G/Agarose column (Invivogen) after column equilibration. Chromatography steps were performed at a flow rate of 0.5ml/min. Next, 20 column volumes of 1X-PBS were used to wash the column. IgG were then eluted with 5ml of 0.1M glycine (pH 2-3, Sigma-Aldrich) and pH was immediately adjusted to 7.5 with 1M Tris. 1X-PBS buffer exchange was achieved using Amicon® Ultra centrifugal filters (Merck Millipore) through a 100-kD membrane according to the manufacturer’s instructions. Quantification of purified IgG was determined using NanoVue Plus microvolume spectrophotometers.

### Pseudovirus production and permissive cell line generation

Pseudotyped vectors were produced by triple transfection of 293T cells as previously described^18^. Briefly, cells were co-transfected with plasmids encoding for lentiviral proteins, a luciferase Firefly reporter, and a plasmid expressing a codon-optimized SARS-CoV-2 Spike gene. Pseudotyped vectors were harvested on day 2 post-transfection. Functional titer (TU) was determined by qPCR after the transduction of a stable HEK 293T-hACE2 cell line. To generate this cell line, HEK 293T cells were transduced at a multiplicity of infection (MOI) 20 with an integrative lentiviral vector expressing the human ACE2 gene under the control of the UBC promoter. Clones were generated by limiting dilution and selected on their permissivity to SARS-CoV-2 S pseudotyped lentiviral vector transduction.

### Pseudoneutralisation Assay

First, serum dilutions were mixed and co-incubated with 300 TU of the pseudotyped vector at room temperature for 30 minutes. Serum and vector were diluted in culture medium or with IVIG batches at a 12mg/mL concentration for IgG. (DMEM-glutamax (Gibco) + 10% FCS (Gibco) + Pen/Strep (Gibco). The mix was then plated in a tissue culture treated black 96-well plate (Costar) with 20 000 HEK 293T-hACE2 cells in suspension. To prepare the suspension, the cell flask was washed with DPBS twice (Gibco) and the cells were individualized with DPBS + 0.1% EDTA (Promega) to preserve hACE2 protein. After 48hrs, the medium was removed and bioluminescence was measured using a Luciferase Assay System (Promega) on an EnSpire plate reader (PerkinElmer). Half maximal inhibitory concentrations IC50 were determined using the Graphpad Prism software 5.

### Funding

MM and GG are supported by the Programme hospitalier de recherche clinique (PHRC: AOR17082,PHRCN 190321), DIM thérapie génique Ile de France and AFPCA (Association Française de la PolyChondrite Atrophiante). D.S. was supported for this work by a Pasteur/APHP interface fellowship. The study was supported by Fondation de France, « Tous unis contre le virus » framework Alliance (Fondation de France, AP-HP, Institut Pasteur) in collaboration with Agence Nationale de la Recherche (ANR Flash COVID19 program), and by the SARS-CoV-2 Program of the Faculty of Medicine from Sorbonne University: ICOViD programs, PI: G.G.).

### Author contributions

A.Mathian, J.F., J.H., Z.A., recruited patients. A.Mathian, M.M, P.Q., T.C., J.F., J.H., Z.A., collected demographic and clinical data. D.S., A.Mohr, L.C., K.D., C.P., P.Q., N.B. prepared the specimens. M.M., D.S., S.Marot, M.A., A.Mohr, F.A, designed and performed experiments. F.A., S.Marot, S.B, D.B, A.G.M, V.C., and P.C. designed, performed and analysed neutralisation assays. S.Mudumba, R.H., C.G. designed and provided the Genalyte system reagents. M.M, S.Marot, F.A. H.Y. and D.S. analysed data. M.M, D.S., F.A. prepared the figures. M.M. H.Y. and G.G. wrote the manuscript draft. M.M., Z.A., G.G. designed the study and reviewed the manuscript.

## Acknowledgments

The authors wish to thank the patients that agreed to participate in this study, doctors and nurses from the endocrinology, metabolism, nutrition and diabetology departments of Institut E3M (Assistance Publique Hôpitaux de Paris (AP-HP), Hôpital Pitié-Salpêtrière, 75013 Paris, France) who participated in this study, and healthy donors.

## References

1. W. Joost Wiersinga, M., PhD; Andrew Rhodes, MD, PhD; Allen C. Cheng, MD, PhD; Sharon J. Peacock, PhD; Hallie C. Prescott, MD, MSc. Pathophysiology, Transmission, Diagnosis, and Treatment of Coronavirus Disease 2019 (COVID-19). JAMA (2020).

2. Grifoni, A., et al. Targets of T Cell Responses to SARS-CoV-2 Coronavirus in Humans with COVID-19 Disease and Unexposed Individuals. Cell 181,1489-1501.e1415 (2020).

3. Le Bert, N., et al. SARS-CoV-2-specific T cell immunity in cases of COVID-19 and SARS, and uninfected controls. Nature (2020).

4. Braun, J., et al. SARS-CoV-2-reactive T cells in healthy donors and patients with COVID-19. Nature (2020).

5. Mateus, J., et al. Selective and cross-reactive SARS-CoV-2 T cell epitopes in unexposed humans. Science (2020).

6. Gelfand, E.W. Intravenous Immune Globulin in Autoimmune and Inflammatory Diseases. New England Journal of Medicine 367, 2015–2025 (2012).

7. Sterlin, D., et al. IgA dominates the early neutralizing antibody response to SARS-CoV-2. *medRxiv*, 2020.2006.2010.20126532 (2020).

8. Premkumar, L., et al. The receptor binding domain of the viral spike protein is an immunodominant and highly specific target of antibodies in SARS-CoV-2 patients. Science immunology 5(2020).

9. Pinto, D., et al. Cross-neutralization of SARS-CoV-2 by a human monoclonal SARS-CoV antibody. Nature 583, 290–295 (2020).

10. Li, F. Structure, Function, and Evolution of Coronavirus Spike Proteins. Annual Review of Virology 3, 237–261 (2016).

11. Lu, R., et al. Genomic characterisation and epidemiology of 2019 novel coronavirus: implications for virus origins and receptor binding. The Lancet 395, 565–574 (2020).

12. Ricke, D. & Malone, R.W. Medical Countermeasures Analysis of 2019-nCoV and Vaccine Risks for Antibody-Dependent Enhancement (ADE). SSRN (2020).

13. Stockman, L.J., Bellamy, R. & Garner, P. SARS: systematic review of treatment effects. PLoS Med 3, e343 (2006).

14. Li, L, et al. Effect of Convalescent Plasma Therapy on Time to Clinical Improvement in Patients With Severe and Life-threatening COVID-19: A Randomized Clinical Trial. JAMA 324, 460–470 (2020).

15. Mudumba, S., et al. Photonic ring resonance is a versatile platform for performing multiplex immunoassays in real time. J Immunol Methods 448, 34–43 (2017).

16. Miyara, M., et al. Detection in whole blood of autoantibodies for the diagnosis of connective tissue diseases in near patient testing condition. PLoS One 13, e0202736 (2018).

17. Sterlin, D., et al. Human IgA binds a diverse array of commensal bacteria. Journal of Experimental Medicine 217(2019).

18. Iglesias, M.C., et al. Lentiviral Vectors Encoding HIV-1 Polyepitopes Induce Broad CTL Responses In Vivo. Molecular Therapy 15, 1203–1210 (2007).

